# The effect of SARS-CoV-2 testing on healthcare seeking behaviour at primary care level: implications for COVID-19 vaccine effectiveness estimates in test-negative design studies

**DOI:** 10.1101/2025.03.31.25324928

**Authors:** Cheyenne C.E. van Hagen, Eric R.A. Vos, Charlotte Lanièce Delaunay, Hester E. de Melker, Esther Kissling, Mirjam J. Knol

## Abstract

**Background:** Diagnostic (self-)testing for SARS-CoV-2 may lead to selection bias in test-negative case-control designs (TND) for COVID-19 vaccine effectiveness (CVE) at primary care level. We investigated whether after the acute phase of the pandemic, (self-)testing among those with an acute respiratory infection (ARI) was associated with healthcare seeking behaviour at primary care level in the general Dutch population.

**Methods:** We pooled questionnaire data from three study rounds (June 2022, November 2022 & April 2023) of the nationwide PIENTER Corona cohort study. Among participants aged 18-91 years, we selected the first self-reported ARI episode, defined as cough, sore throat, dyspnoea and/or coryza, since March 2022. We performed log-binomial regression analyses adjusted for age, sex, educational level and comorbidities to assess associations between COVID-19 vaccination, SARS-CoV-2 (self-)testing and general practitioner (GP) consultation, and between GP consultation and prior (self-)test result.

**Results:** Among 3152 participants with an ARI episode, vaccinated (vs unvaccinated) participants more often (self-)tested (adjusted RR [95% CI]: 1.07 [1.04–1.11]) or consulted a GP (1.57 [1.21–2.09]). (Self-)test result overall was not associated with GP consultation (0.86 [0.69–1.08]). Vaccination-stratified analyses showed vaccinated individuals were less likely to consult the GP after a positive (self-) test (0.62 [0.49–0.79]), while unvaccinated were more likely to (2.00 [1.08–3.51]).

**Conclusions:** In this Dutch population-based cohort, GP consultation between May 2022 and March 2023 was differential by (self-)test result and vaccination status, indicating potential selection bias in TND CVE estimates from testing before GP consultation. More research to quantify this bias in various settings is needed.

## Introduction

Vaccination has been a key tool in controlling the COVID-19 pandemic by reducing the risk of mortality and severe disease after SARS-CoV-2 infection (1). To assess the performance of vaccines, COVID-19 vaccine effectiveness (CVE) estimates are a crucial source of information. CVE studies at primary care/outpatient level generally provide information on COVID-19 vaccines performance against relatively mild disease. Knowledge on this topic is important to guide public health policy.

The test-negative design (TND) study has been frequently used for influenza vaccine effectiveness at primary care level and in other settings, and has been used for CVE as well (2). Patients presenting to a general practitioner (GP) that meet a specified clinical case definition are swabbed in the nasopharynx. Those testing positive for SARS-CoV-2 are designated as cases and those testing negative as controls (3). By design, an advantage of TND studies is that it reduces the risk of bias through healthcare seeking behaviour (HSB) as only people with similar HSB, those consulting the GP, are included (4, 5). Prerequisites of TND studies towards vaccine effectiveness include that controls are representative of the source population from which the cases arise in terms of vaccination.

Traditionally, both patients and practitioners are blind to the pathogen causing the symptoms at time of consultation. However, this changed during the COVID-19 pandemic when large-scale testing for SARS-CoV-2 was implemented, first at municipal health services and later through rapid antigen self-tests. With testing for SARS-CoV-2 widely available, the general population can obtain a COVID-19 diagnosis at home, compromising the blinded aspect of TND CVE studies and potentially causing selection bias (6). Since both rapid antigen self-tests and SARS-CoV-2 tests administered by a third party (e.g. municipal health services) can reveal a COVID-19 diagnosis before GP consultation, we refer to all SARS-CoV-2 diagnostic testing (i.e. self-administered rapid antigen tests and third-party-administered PCR or rapid antigen tests) preceding GP consultation as “self-tests”. The testing behaviour of a population is expected to vary over time and by age and other characteristics, as well as according to government guidelines and recommendations. The availability of self-tests may bias TND CVE estimates at primary care level if GP consultation is related to self-testing and vaccination status (6).

Here, we aimed to assess whether self-testing may result in selection bias in TND CVE estimates. To that regard we investigated to what extend the use and result of self-tests for acute respiratory infections (ARI) in the general Dutch population is associated with ARI-related HSB at primary care level between March 2022 and May 2023, and whether other factors such as vaccination status, age and underlying conditions influence this association.

## Methods

### Study design & population

We used data from PIENTER Corona (PICO), a prospective nationwide population-based seroepidemiological cohort study aimed at monitoring humoral immunity against SARS-CoV-2 in the Netherlands (7, 8). The first round (PICO1) was conducted in April 2020. Initially, participants were sampled from the 2016/2017 established PIENTER3 serosurvey cohort which included Dutch residents randomly selected via a two-stage cluster design (7, 9). To enhance countrywide geographical coverage and increase and maintain power, the study population was supplemented twice (in June 2020 - PICO2, and November 2021 - PICO6) with two additional random samples of persons from the Dutch population registry (8, 10). This yielded an overall study population of 11 159 participants aged 1–92 years.

Thirteen PICO study rounds have been conducted until the study’s end in late 2024 (Supplementary file – Figure S1). In each study round, participants submitted a finger-prick blood sample in a microtainer by mail and completed a questionnaire on sociodemographic factors, health status, COVID-19 vaccination, and SARS-CoV-2-related symptoms. Since PICO4 (February 2021), questions on vaccination, episodes of ARI, self-testing, and healthcare consultations have been included in the questionnaire (Supplementary file – Figure S2).

For this analysis we included only participants ≥18 years because of differences in vaccination rollout and testing behaviour between children and adults. We used data from PICO8–PICO10 with the majority of inclusions covering June 2022, November 2022 and April 2023, respectively. This period took place after the acute phase of the pandemic with most restrictive measures lifted. From April 2022 onwards, the Dutch government still advised self-testing when experiencing COVID-like symptoms and PCR testing at municipal health services was only recommended for certain (risk) groups (11). In March 2023, the advice on self-testing was discontinued and PCR testing facilities closed (12).

### Statistical analysis

Statistical analyses followed two hypothesized ways, as described by Lanièce Delaunay et al, that self-testing could introduce selection bias in TND CVE estimates (Figure 1) (6). Firstly, if COVID-19 vaccination status influences self-testing independently from HSB and the result of the self-test influences one’s decision to consult the GP, selecting on GP consultation creates an open non-causal path between COVID-19 vaccination and SARS-CoV-2 infection (13). Secondly, even in the absence of an association between COVID-19 vaccination and self-testing, vaccination could affect GP consultation and act as an effect modifier of the association between self-test result and GP consultation if among those using self-tests, consulting a GP is differential by self-test result and vaccination status and other factors such as age and underlying health conditions.

**Figure 1:**
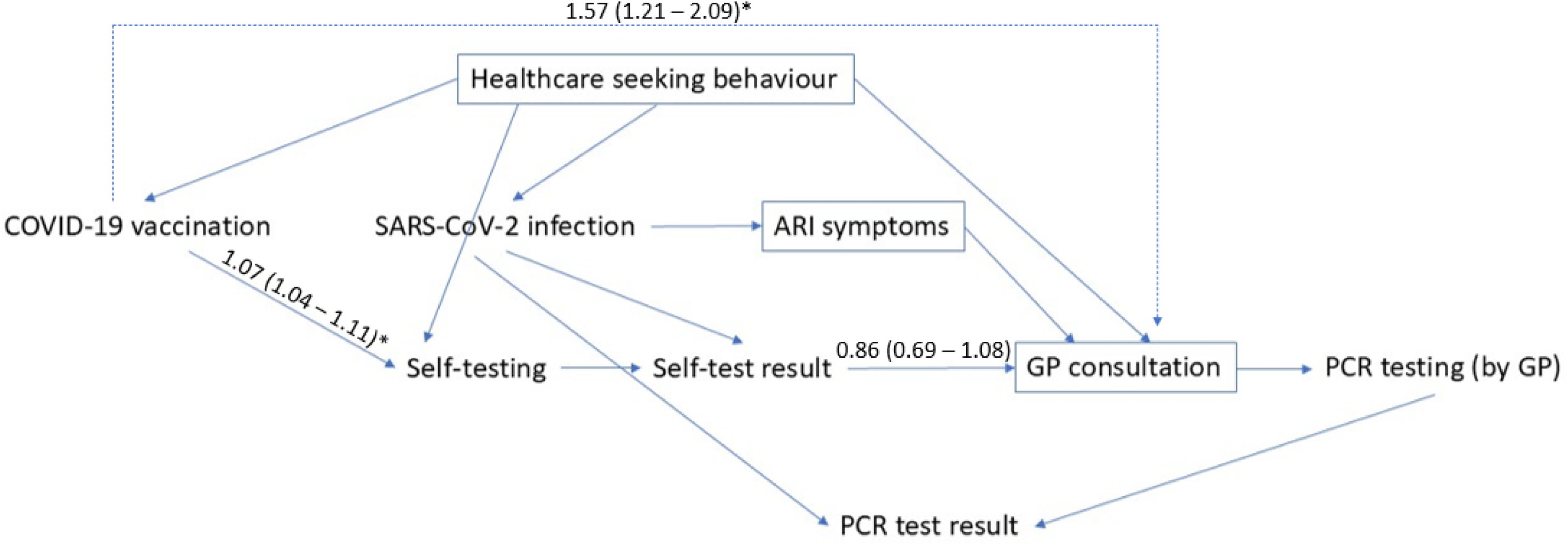
Directed Acyclic Graph (DAG) showing the two potential routes to biased CVE estimates (based and elaborated on Lanièce Delaunay et al (6), figure 2). Firstly, if COVID-19 vaccination is directly associated with self-testing and the self-test result is associated with GP consultation, there is an open non-causal path between COVID-19 vaccination and SARS-CoV-2 infection in a TND study (COVID-19 vaccination → Self-testing → Self-test result ← SARS-CoV-2 infection) as a result of selecting on GP consultation, a descendent of the collider self-test result. Secondly, if COVID-19 vaccination is associated with GP consultation, a causal structure occurs in which COVID-19 vaccination might act as an effect modifier of the association between self-test result and GP consultation (19). Adjusted risk ratios of associations between COVID-19 vaccination and GP consultation, COVID-19 vaccination and self-testing and self-test result and GP consultation are depicted at corresponding arrows in the DAG.

We pooled the data from PICO8–10 and selected participants with ARI. ARI was defined as having at least one of cough, sore throat, shortness of breath or coryza. Participants could report up to three symptom episodes per study round. If multiple episodes matched the ARI case definition, the first reported episode was selected.

For all analyses, complete case selection was conducted for variables age, sex, educational level, underlying conditions, COVID-19 vaccination status, self-testing and GP consultation. Age was calculated using date of birth from the Dutch population registry and questionnaire completion date and categorized into 18-35, 36-64 and ≥65 years. Sex (male/female) was retrieved from the Dutch population registry. Educational level was classified as low (no education or primary education), intermediate (secondary school or vocational training), or high (higher professional/academic education) based on self-reported questionnaire data. The presence of underlying conditions was also based on self-reported questionnaire data and defined as eligibility for flu vaccination in the Netherlands (yes/no). This included presence of at least one of asthma, diabetes, asplenia, cardiovascular disease, immune deficiency, cancer (currently treated and currently untreated), liver disease, lung disease, neurological disease, kidney disease and organ or bone marrow transplantation. COVID-19 vaccination status was based on the Dutch COVID-19 vaccination registry (CIMS) (14) and complemented with self-reported questionnaire data if CIMS did not contain a vaccination record (i.e. when unvaccinated or not consented to vaccination registration). Participants with at least one COVID-19 vaccine dose prior to ARI onset were considered vaccinated. Those vaccinated were subclassified into number of doses received (1–2; 3 or ≥4 doses) and time since last vaccination (0–90; 91–180; 180+ days). GP consultation (yes/no), self-testing (yes/no) and self-test result (positive/negative) were identified from self-reported questionnaire data. Self-tests included self-administered rapid antigen tests and third-party-administered PCR/rapid antigen tests.

In descriptive analyses the proportion of the population taking a self-test when having ARI symptoms was determined. Among the study population with ARI symptoms (Figure 2, light blue circle), participant characteristics were presented overall and stratified by vaccination status for ARI onset date, age, sex, ethnic background, educational level, presence of underlying conditions, COVID-19 vaccination status, self-testing and result and GP consultation. Additionally, among those self-testing for ARI symptoms (Figure 2, dark blue circle), similar relevant descriptive statistics were presented overall and stratified by self-test result.

**Figure 2:**
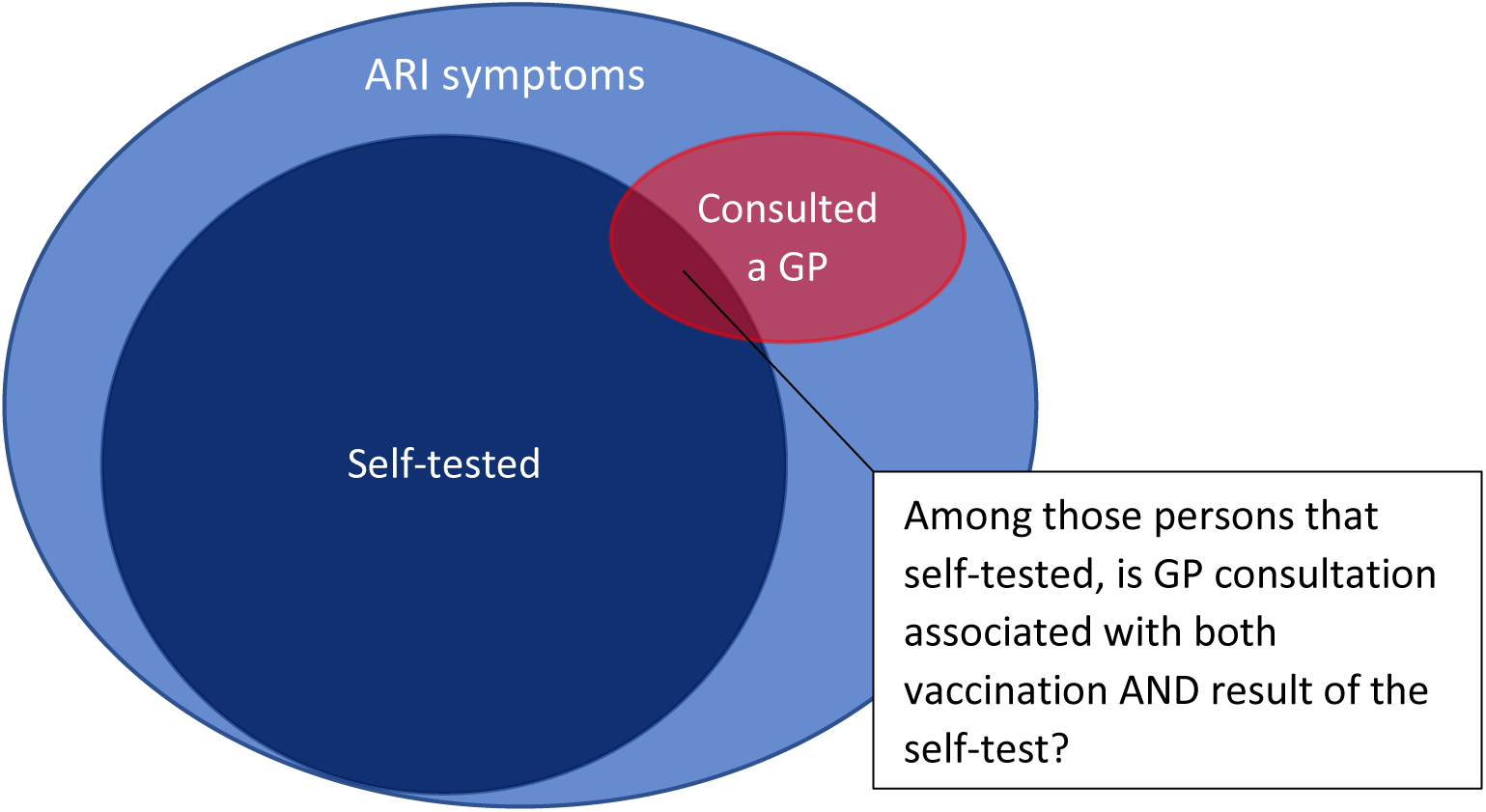
Population with ARI symptoms self-testing and consulting a GP.

Log binomial regression models were fitted to assess associations between COVID-19 vaccination status and self-testing and GP consultation, respectively, and adjusted for age, sex, educational level and presence of underlying conditions. Crude (RR) and adjusted risk ratios (aRR) and 95% confidence intervals (95% CI) of self-testing and GP consulting were presented. In addition, among the population with ARI symptoms that took a self-test, log binomial regression models were fitted to assess whether consulting the GP after taking a self-test (Figure 2, dark blue + red circle) depends on the self-test result, and adjusted for age, sex, educational level and presence of underlying conditions. This analysis was repeated stratified by age group, presence of underlying conditions and COVID-19 vaccination status to determine whether the association differs between these groups. Secondarily, for those vaccinated, stratification by number of COVID-19 vaccinations and time since last COVID-19 vaccination was performed.

Data cleaning was conducted in SAS 94 M7 English and statistical analyses were performed using R version 4.4.1.

## Results

### Characteristics of ARI participants

From a total of 3236 participants with ARI symptoms (Supplementary file – Table S1), complete case analysis was conducted with 3152 (97%) participants (Table 1). Median onset date of first ARI episode was 20 July 2022 (IQR 20 May 2022 – 23 October 2022). ARI symptoms most often reported included cough (66%), coryza (63%) or sore throat (62%) and 15% reported shortness of breath. Participants had a median age of 53 years (IQR 39–67 years) and 64% was female. About a quarter of the study population (27%) had an underlying condition, with cardiovascular disease most often reported (17% of study population). During the ARI episode, 87% of participants took a self-test, with numbers varying by age group (18–35y: 79%; 36–64y: 89%; ≥65y: 89%). Of those that self-tested, 84% took a self-administered rapid antigen test (as opposed to a third-party administered PCR or rapid antigen test). One in ten participants consulted the GP for their ARI symptoms, which varied by age group (18– 35y: 7%; 36–64y: 8%; ≥65y: 16%). At ARI onset, 69% of the study population had received at least one COVID-19 vaccination, of which the majority had received 3 doses (52%) and received their last dose >90 days before ARI onset (72%). Those vaccinated for COVID-19 (vs unvaccinated) were generally older (median [IQR] age: 57 [42–70] vs 47 [35–59] years), more often reported underlying conditions (29% vs 23%), more often self-tested (89% vs 82%) and more often consulted a GP (11% vs 6%) (Table 1).

**Table 1.**
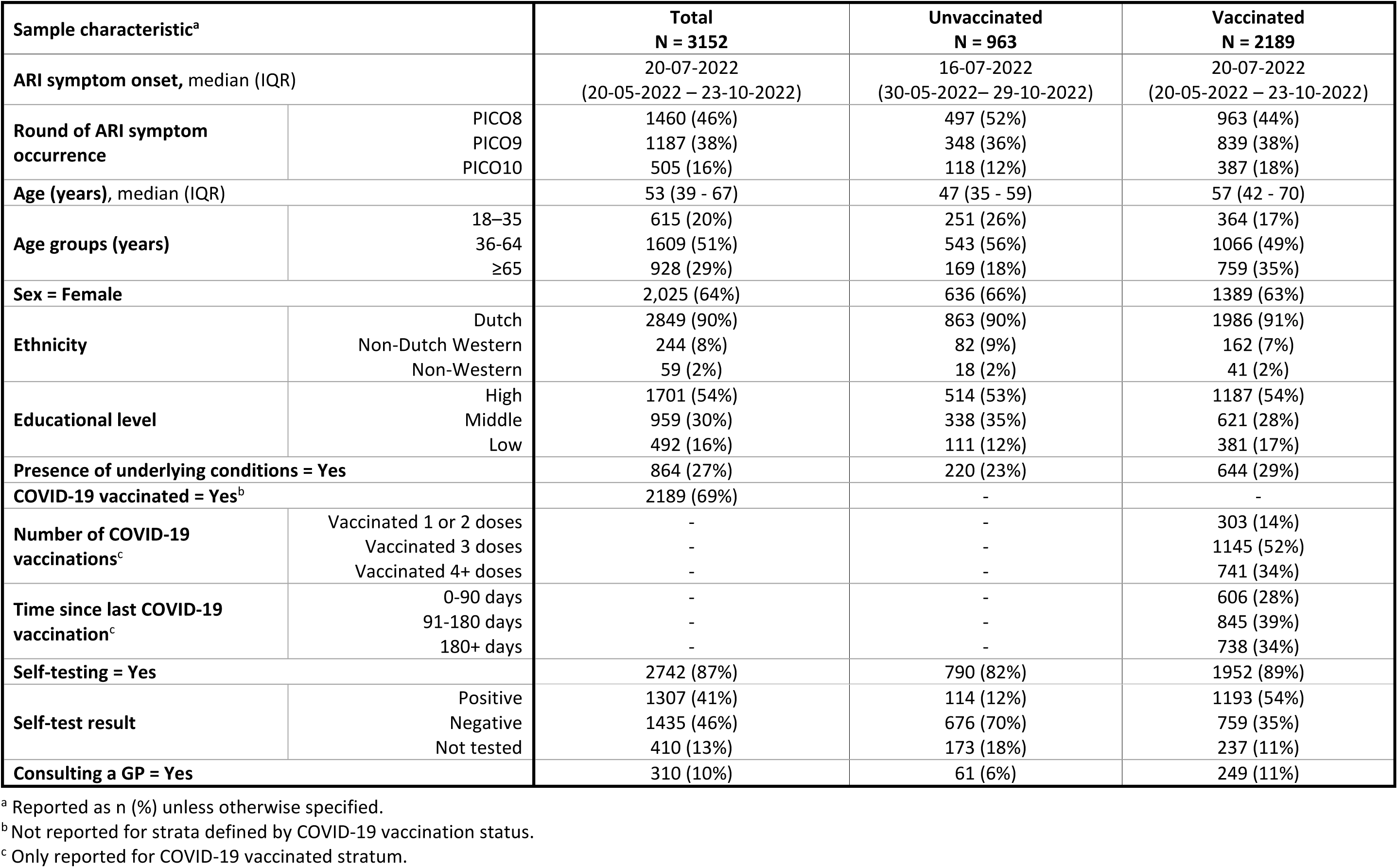
Characteristics of participants with ARI symptoms, stratified by COVID-19 vaccination status.

#### COVID-19 vaccination, self-testing and GP consultation

COVID-19 vaccination was statistically significantly associated with both self-testing (aRR [95% CI]: 1.07 [1.04–1.11]) and GP consultation (aRR [95% CI]: 1.57 [1.21–2.09]) (Table 2).

**Table 2.** The association between COVID-19 vaccination status and self-testing and GP consultation.

|  |  | **COVID-19 vaccination status** |  | **RR**<br>**(95% CI)** | **aRR**<br>**(95% CI)** |
| --- | --- | --- | --- | --- | --- |
|  |  | Vaccinated | Unvaccinated |  |  |
| **Self-testing** | Yes | 1952 | 790 | 1.09 | 1.07 |
|  | No | 237 | 173 | (1.05 – 1.12) | (1.04 – 1.11) |
| **GP consultation** | Yes | 249 | 61 | 1.80 | 1.57 |
|  | No | 1940 | 902 | (1.38 – 2.37) | (1.21 – 2.09) |

### Characteristics of ARI participants that self-tested

Among 2742 participants that self-tested for their ARI symptoms, median age was 54 years (IQR 40– 67 years) and 65% was female (Table 3). About one in four (28%) had an underlying condition and 71% was vaccinated. A minority of self-testing participants consulted the GP for the ARI symptoms (10%). Self-test result was nearly evenly distributed with 48% of those that self-tested reporting a positive test result. Those with a positive self-test (vs negative self-test) were generally older (median [IQR] age: 60 [46–72] vs 49 [37–62] years) and less often female (62% vs 68%), more often reported underlying conditions (32% vs 25%) and were more often vaccinated (91% vs 53%) (Table 3).

**Table 3.** Characteristics of participants with ARI symptoms that self-tested, stratified by self-test result.

| Sample characteristic<sup>a</sup> |  | Total<br>N = 2742 | Positive self-test<br>N = 1435 | Negative self-test<br>N = 1307 |
| --- | --- | --- | --- | --- |
| Age (years), median (IQR) |  | 54 (40 - 67) | 49 (37 - 62) | 60 (46 - 72) |
| Age groups (years) | 18–35 | 484 (18%) | 312 (22%) | 172 (13%) |
|  | 36–64 | 1436 (52%) | 820 (57%) | 616 (47%) |
|  | ≥65 | 822 (30%) | 303 (21%) | 519 (40%) |
| Sex = Female |  | 1792 (65 %) | 976 (68%) | 816 (62%) |
| Ethnicity | Dutch | 2487 (91%) | 1294 (90%) | 1193 (91%) |
|  | Non-Dutch Western | 206 (8%) | 110 (8%) | 96 (7%) |
|  | Non-Western | 49 (2%) | 31 (2%) | 18 (1%) |
| Educational level | High | 1478 (54%) | 835 (58%) | 643 (49%) |
|  | Middle | 829 (30%) | 426 (30%) | 403 (31%) |
|  | Low | 435 (16%) | 174 (12%) | 261 (20%) |
| Presence of underlying conditions = Yes |  | 768 (28 %) | 352 (25%) | 416 (32%) |
| COVID-19 vaccinated = Yes |  | 1952 (71 %) | 759 (53%) | 1193 (91%) |
| Number of COVID-19<br>vaccinations<sup>b</sup> | Vaccinated 1 or 2 doses | 246 (9%) | 102 (7%) | 144 (11%) |
|  | Vaccinated 3 doses | 1036 (38%) | 449 (31%) | 587 (45%) |
|  | Vaccinated 4+ doses | 670 (24%) | 208 (14%) | 462 (35%) |
| Time since last COVID-19<br>vaccination<sup>b</sup> | 0-90 days | 561 (20%) | 200 (14%) | 361 (28%) |
|  | 91-180 days | 747 (27%) | 299 (21%) | 448 (34%) |
|  | 180+ days | 644 (23%) | 260 (18%) | 384 (29%) |
| Self-test result<sup>c</sup> | Positive | 1307 (48%) | - | - |
|  | Negative | 1435 (52%) | - | - |
| Consulting a GP = Yes |  | 280 (10%) | 147 (10%) | 133 (10%) |

<sup>a</sup> Reported as n (%) unless otherwise specified.
<sup>b</sup> Reported in percentage of total (including unvaccinated, though not shown).
<sup>c</sup> Not reported for strata defined by self-test result.

#### Self-test result and GP consultation

Overall, no statistically significant association was observed between self-test result and GP consultation where the risk of consulting a GP after a positive self-test result was 14% lower (0.86 [95% CI: 0.69–1.08]) than after a negative self-test result (Table 4). Stratification by age groups also did not show statistically significant associations between self-test result and GP consultation, though point estimates increased with age (aRR [95% CI] 18–35y: 0.68 [0.33–1.27]; 36–64y: 0.79 [0.54–1.12]; ≥65y: 0.99 [0.71–1.40]). Similarly, among those with and without underlying conditions, non-statistically significant associations were observed between self-test result and GP consultation (respective aRR [95% CI]: 1.01 [0.73–1.41] and 0.76 [0.55–1.03]).

**Table 4:** The association between self-test result and GP consultation among persons with ARI symptoms that self-tested.

|  |  | Self-test result | GP consultation |  | RR (95% CI) | aRR (95% CI) |
| --- | --- | --- | --- | --- | --- | --- |
|  |  |  | Yes | No |  |  |
| **Overall** |  | Positive | 133 | 1174 | 0.99 | 0.86 |
|  |  | Negative | 147 | 1288 | (0.79 – 1.24) | (0.69 – 1.08) |
| **Age group<sup>1</sup>** | 18-35 years | Positive | 11 | 161 | 0.69 | 0.68 |
|  |  | Negative | 29 | 283 | (0.34 – 1.30) | (0.33 – 1.27) |
|  | 36-64 years | Positive | 44 | 572 | 0.81 | 0.79 |
|  |  | Negative | 72 | 748 | (0.56 – 1.16) | (0.54 – 1.12) |
|  | 65+ years | Positive | 78 | 441 | 0.99 | 0.99 |
|  |  | Negative | 46 | 257 | (0.71 – 1.40) | (0.71 – 1.40) |
| **Underlying conditions<sup>2</sup>** | Yes | Positive | 69 | 347 | 1.06 | 1.01 |
|  |  | Negative | 55 | 297 | (0.77 – 1.47) | (0.73 – 1.41) |
|  | No | Positive | 64 | 827 | 0.85 | 0.76 |
|  |  | Negative | 92 | 991 | (0.62 – 1.15) | (0.55 – 1.03) |
| **COVID-19 vaccination status** | Vaccinated | Positive | 119 | 1074 | 0.68 | 0.62 |
|  |  | Negative | 111 | 648 | (0.54 – 0.87) | (0.49 – 0.79) |
|  | Unvaccinated | Positive | 14 | 100 | 2.31 | 2.00 |
|  |  | Negative | 36 | 640 | (1.24 – 4.04) | (1.08 – 3.51) |
| **Number of COVID-19 vaccinations** | Vaccinated 1-2 doses | Positive | 15 | 129 | 1.18 | 1.13 |
|  |  | Negative | 9 | 93 | (0.55 – 2.71) | (0.54 – 2.55) |
|  | Vaccinated 3 doses | Positive | 36 | 551 | 0.44 | 0.41 |
|  |  | Negative | 62 | 387 | (0.30 – 0.65) | (0.28 – 0.61) |
|  | Vaccinated 4+ doses | Positive | 68 | 394 | 0.77 | 0.74 |
|  |  | Negative | 40 | 168 | (0.54 – 1.10) | (0.52 – 1.06) |
| **Time since last COVID-19 vaccination** | 0-90 days | Positive | 39 | 322 | 0.68 | 0.65 |
|  |  | Negative | 32 | 168 | (0.44 – 1.05) | (0.42 – 1.01) |
|  | 91-180 days | Positive | 43 | 405 | 0.53 | 0.48 |
|  |  | Negative | 54 | 245 | (0.36 – 0.77) | (0.33 – 0.71) |
|  | 180+ days | Positive | 37 | 347 | 1.00 | 0.82 |
|  |  | Negative | 25 | 235 | (0.62 – 1.64) | (0.51 – 1.33) |

<sup>1</sup> Age-stratified analyses were only adjusted for sex, educational level and presence of underlying conditions.
<sup>2</sup> Underlying condition-stratified analyses were only adjusted for age, sex and educational level.

After stratification by COVID-19 vaccination status, aRRs of GP consultation after a positive self-test result were significant with different effect directions. The risk of consulting a GP after a positive self-test result compared to a negative self-test result, was 0.62 [95% CI: 0.49–0.79] among the vaccinated and 2.00 [95% CI: 1.08–3.51] among the unvaccinated. Among those vaccinated, further stratification by the number of doses received and time since last vaccination showed significant negative associations between a positive self-test and GP consultation only among those vaccinated with 3 doses (aRR [95% CI]: 0.41 [0.28–0.61]) and those last vaccinated 91–180 days ago (aRR [95% CI]: 0.48 [0.33–0.71]). Other point estimates among these strata did not show consistent increasing or decreasing patterns.

### Selection bias in TND CVE estimates

Linking our results to the hypotheses of Lanièce Delaunay et al, shows proof for one of the two ways of bias in CVE estimates (6).

First of all, self-testing was associated with COVID-19 vaccination status, but GP consultation after self-testing was overall not associated with the self-test result (Figure 1). Therefore, we cannot establish the occurrence of bias through the non-causal path COVID-19 vaccination ◊ Self-testing ◊ Self-test result ⇓ SARS-CoV-2 infection (Figure 1).

Secondly, COVID-19 vaccination was significantly associated with GP consultation and the risk of GP consultation after a positive compared to a negative self-test result was significantly lower among vaccinated and higher among unvaccinated individuals (Figure 1). Thus, COVID-19 vaccination was an effect modifier of the association between self-test result and GP consultation. Our findings within this Dutch study therefore support the second hypothesized way of bias and suggests that selection bias in TND studies from self-testing is a valid point of concern.

## Discussion

In this nationwide cohort study among the general Dutch population, we assessed whether self-testing could bias TND CVE estimates by investigating associations between ARI-related self-testing and HSB at primary care level after the acute phase of the pandemic. Our findings show that between March 2022 and May 2023, COVID-19 vaccination status was statistically significantly associated with both self-testing and GP consultation. Overall, people with a positive self-test were less likely to consult a GP for ARI symptoms, but this was not statistically significant. Increasing point estimates with increasing age suggest that older people were more likely to consult the GP after a positive test than younger people. In contrast to unvaccinated persons, vaccinated participants with a positive self-test were statistically significantly less likely to consult the GP than those with a negative self-test result, which may potentially lead to selection bias in TND CVE.

TND studies generally address confounding by unmeasured HSB when estimating vaccine effectiveness against infection (4, 5). To that regard several important assumptions are made (4). One of these assumptions is that all patients consulting the GP have the same HSB which might be invalid for COVID-19 where the blinded aspect of TND studies is compromised as a result of self-testing. Self-testing can potentially cause two ways leading to biased CVE estimates resulting from selection bias as described by Lanièce Delaunay et al (6). Our findings within this Dutch study proved only the second pathway where GP consultation was differential by self-test result and COVID-19 vaccination status. In our case, this could lead to an overestimation of CVE if not weighted for GP consultation probabilities of all combined strata of vaccination status and self-test result (6). Although GP consultation overall was not significantly associated with self-test result, the first pathway may still be present within subgroups. Younger people and those without underlying conditions seemed less likely to consult the GP after a positive test than older people and those with underlying conditions respectively, though also no significant results were found.

Though some publications address the topic of biased CVE estimates as a result of non-equal HSB, very few publications discuss the issue of self-testing in a similar context (4, 5, 15–18). According to Dean et al, the blinded aspect of TND studies in which study inclusion does not depend on the obtained test result is crucial to prevent selection bias, which makes retrospective designs prone to biased CVE estimates (15). Shi et al hypothesize that widespread and regular self-testing is one of the challenges in TND studies for COVID-19 which mostly influences the recruitment of test-negative participants (4). Lewnard et al and Vandenbroucke et al elaborate that there is large variation in reasons for testing which is important to account for in the analysis to prevent biased CVE (16, 17). Lanièce Delaunay et al were the first to elaborately address the potential selection bias in TND studies due to self-testing and performed a simulation study to investigate its impact on CVE estimates (6). To our knowledge, our study is the first to prove selection bias in TND CVE studies from self-testing.

A great strength of our study is that we use a large population-based cohort to assess HSB in the general population. However, our study also has some limitations. First, participation in our study might be associated with HSB and our study population might be more motivated to self-test or seek care when ill. Though this might have an influence on the proportion that self-tests, we don’t expect this to largely influence our results regarding selection bias since this accounts for all participants, whether they tested positive or negative. Secondly, because of low GP consultation proportions, we pooled data from different study rounds to generate sufficient power. Testing behaviour varied over time in our data as self-testing rates among ARI participants fluctuated somewhat between study round (PICO8: 87%; PICO9 91%; PICO10: 76%), though numbers were consistently high. According to the simulation study of Lanièce Delaunay et al, a 20-percentage point increase in overall self-testing rate could increase the mean bias in estimated VE with 11-17 points. Therefore, the fluctuating self-testing rates between study rounds might impact the magnitude of bias in our study, but not its direction. Thirdly, we did not determine whether self-tests were taken prior to GP consultation as this data was unavailable in PICO8 and 9. Therefore, post-consultation self-testing might be present in our data, though we assume that self-testing is mostly performed before consulting the GP. An extra question was added to the PICO10 questionnaire to identify when a self-test was taken in relation to GP consultation (Supplementary file – Figure S2). In PICO10, 114 participants self-tested and consulted the GP of which 6% took a self-test after consulting the GP. With the majority of self-tests (94%) taken prior to GP consultation, we do not expect post-consultation self-testing to largely influence our results. Finally, to estimate the magnitude of bias in TND CVE estimates, we attempted to estimate CVE with and without bias using a nested test negative study with serologically identified infections. However, due to the low proportion of GP consultations we were not able to conduct this analysis with sufficient precision and it is therefore not presented here. While the exact bias magnitude cannot be determined, our results, when compared to the simulation study results of Lanièce Delaunay et al (6), suggest a greater bias based on the high proportion of self-testing in our population (87% in our study), but a relatively small influence on bias based on the low difference between the probability of self-testing among vaccinated compared to unvaccinated (1.07 in our study).

In this Dutch population-based cohort between May 2022 and March 2023, GP consultation was differential by self-test result and COVID-19 vaccination status, indicating potential selection bias in TND CVE estimates due to self-testing. Our findings suggest that this may cause an overestimation of CVE, though this may be context specific. Further research with a larger study population and in different settings is needed to further explore and quantify this bias.

## Ethics approval

The PICO study was conducted in accordance with the principles of the Declaration of Helsinki and the study protocol was approved by the Medical Ethics Committee MEC-U, the Netherlands (Clinical Trial Registration NTR8473). All participants provided written informed consent.

## Data availability

The data that support the findings of this study can be requested via the PICO study website (https://www.rivm.nl/en/pienter-corona-study/information-for-researchers). Restrictions may apply to the availability of these data.

## Acknowledgements

We would like to thank our colleagues from the National Institute of Public Health and Environment (RIVM), Centre of Infectious Disease Control for their contributions regarding logistics, laboratory analyses, methodological insights and manuscript reviewing.

