## Supplementary file for "The effect of SARS-CoV-2 testing on healthcare seeking behaviour at primary care level: implications for COVID-19 vaccine effectiveness estimates in test-negative design studies"

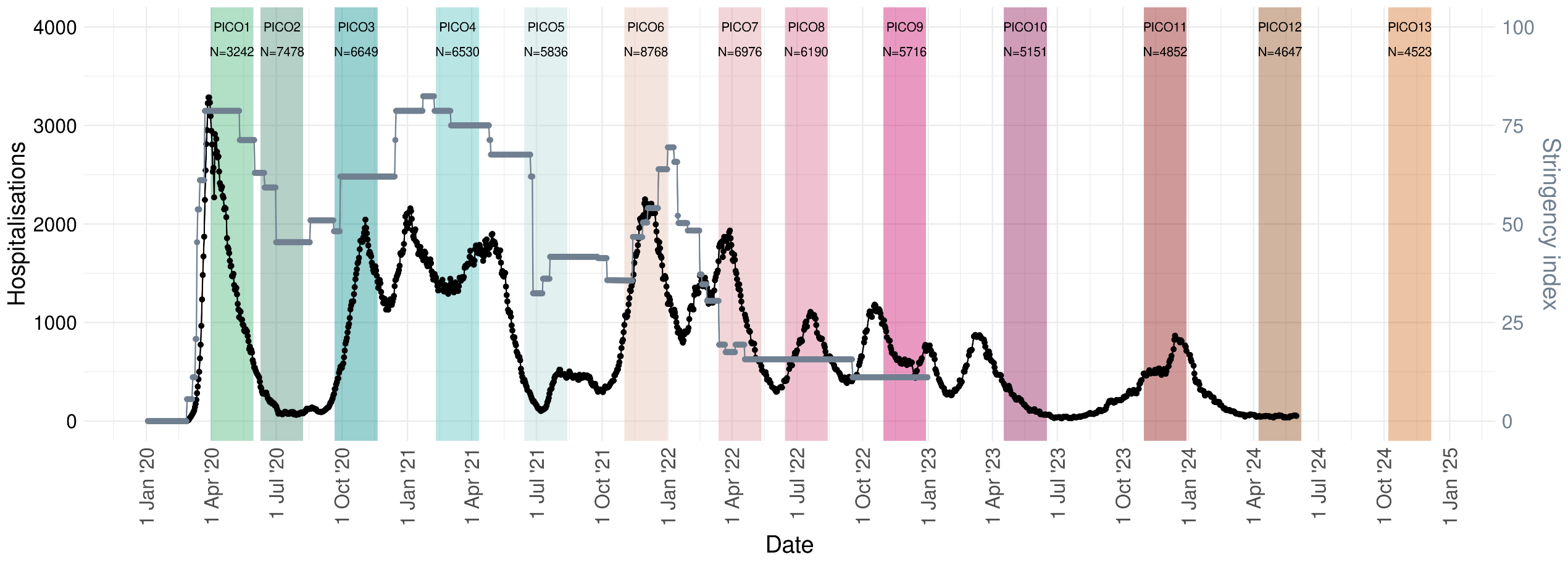
**Figure S1:** Timeline of the PIENTER Corona (PICO) study rounds relative to COVID-19 hospitalisations^1^ (black line, left y-axis) and stringency of restrictive measures^2^ (grey line, right y-axis) in the Netherlands between January 2020 and August 2024.

^1^ Data on hospitalisations are open source and were downloaded from the National Coordination Centre for Patient Distribution (LCPS) (via <https://lcps.nu/datafeed>).

^2^ Stringency index indicates the strictness of government policies and ranges from 0 to 100 (strictest). Stringency index data are open source and were downloaded from Our World in Data (via <https://ourworldindata.org/covid-stringency-index>).

**Figure S2**. Questions asked around ARI, self-testing and GP consultations in PICO since round 4 (PICO4). Questions in green were added in the tenth PICO round (PICO10).

- Did you have any symptoms possibly related to COVID since the last round of the study?
  - If yes:
  - Which symptoms: fever/chills, general malaise, coughing, sore throat, runny nose, shortness of breath, diarrhea, nausea/vomiting, headache, irritability/confusion, muscle ache, pain when breathing, stomach ache, joint pain, loss/change of smell and/or taste, extreme fatigue, pain behind the eyes, dizziness and other symptoms.
  - When did the symptoms start / end?
  - Did you take one or more COVID tests?
  - If yes:
    - What kind of test(s): PCR test, self-administered antigen rapid test or antigen rapid test at a testing facility?
    - Was the result of any of the tests positive for COVID?
  - If yes
    - What was the date the (first) positive test was taken?
    - For which test(s) did you have a positive result: PCR test, self-administered antigen rapid test or antigen rapid test at a testing facility?
  - Did you consult your GP for the symptoms?
  - If yes (and if one or more COVID tests were taken):
    - When did you take the COVID test(s) in relation to your GP consult: a positive test prior to the GP consult, a negative test prior to the GP consult, during the GP consult (by the GP) or after the GP consult.
  - Were you admitted to the hospital for the symptoms?

**Table S1**. Characteristics of participants with ARI symptoms in response and complete case sample.

|  | | **Response sample N=3236** | **Complete case sample**  **N=3152** |
| --- | --- | --- | --- |
| **ARI symptom onset** | Median (IQR) | 18-07-2022  (20-05-2022 – 23-10-2022) | 20-07-2022  (20-05-2022 – 23-10-2022) |
| **Round of ARI symptom occurrence** | PICO8 | 1,511 (47%) | 1,460 (46%) |
|  | PICO9 | 1,211 (37%) | 1,187 (38%) |
|  | PICO10 | 514 (16%) | 505 (16%) |
| **Age** | Median (IQR) | 54 (39 - 67) | 53 (39 - 67) |
| **Age groups (years)** | 18–35 | 628 (19%) | 615 (20%) |
|  | 36-64 | 1,639 (51%) | 1,609 (51%) |
|  | ≥65 | 969 (30%) | 928 (29%) |
| **Sex** | Female | 2,076 (64%) | 2,025 (64%) |
| **Ethnicity** | Dutch | 2,925 (90%) | 2,849 (90%) |
|  | Non-Dutch Western | 248 (8%) | 244 (8%) |
|  | Non-Western | 63 (2%) | 59 (2%) |
| **Educational level** | High | 1,730 (53%) | 1,701 (54%) |
|  | Middle | 986 (30%) | 959 (30%) |
|  | Low | 516 (16%) | 492 (16%) |
|  | Missing | 4 (0%) | 0 |
| **Presence of underlying conditions** | Yes | 885 (27%) | 864 (27%) |
|  | No | 2,323 (72%) | 2,288 (73%) |
|  | Missing | 28 (1%) | 0 |
| **COVID-19 vaccination status** | Unvaccinated | 1,003 (31%) | 963 (31%) |
|  | Vaccinated | 2,232 (69%) | 2,189 (69%) |
|  | Missing | 1 (0%) | 0 |
| **Number of COVID-19 vaccinations** | Unvaccinated | 1,003 (31%) | 963 (31%) |
|  | Vaccinated 1 or 2 dose | 307 (9%) | 303 (10%) |
|  | Vaccinated 3 doses | 1,163 (36%) | 1,145 (36%) |
|  | Vaccinated 4+ doses | 762 (24%) | 741 (24%) |
|  | Missing | 1 (0%) | 0 |
| **Time since last COVID-19 vaccination** | Unvaccinated | 1,003 (31%) | 963 (31%) |
|  | 0-90 days | 622 (19%) | 606 (19%) |
|  | 91-180 days | 863 (27%) | 845 (27%) |
|  | 180+ days | 747 (23%) | 738 (23%) |
|  | Missing | 1 (0%) | 0 |
| **Self-testing** | Yes | 2,815 (87%) | 2,742 (87%) |
|  | No | 412 (13%) | 410 (13%) |
|  | Missing | 9 (0%) | 0 |
| **Self-test result** | Positive | 1,324 (41%) | 1,307 (41%) |
|  | Negative | 1,447 (45%) | 1,435 (46%) |
|  | Not tested | 412 (13%) | 410 (13%) |
|  | Missing | 53 (2%) | 0 |
| **Consulting a GP** | Yes | 320 (10%) | 310 (10%) |
|  | No | 2,908 (90%) | 2,842 (90%) |
|  | Missing | 8 (0%) | 0 |
